# Assessing ChatGPT’s Performance in Delineating Uveitis: An analysis of responses to real-world case presentations

**DOI:** 10.1101/2025.07.05.25330926

**Authors:** Muhammad Sohail Halim, Aly Hamza Khowaja, Zoha Zahid Fazal, Tanya Jain, Kholood Janjua, Ammar Aamir Khan, Anh Ngoc Tram Tran, Yasir J Sepah

## Abstract

**Background:** In the world of Artificial Intelligence (AI), Generative Pretrained Transformer-3 (GPT-3), has gained significant popularity for its demonstrated potential in medical education and diagnostics.

**Rationale:** While AI has shown promising results in healthcare thus far, its understanding of ocular urgencies, particularly uveitis, demands a focused investigation.

**Methods:** This study explored the application of ChatGPT, a language model derived from GPT-3, in delineating uveitis based on patient presentations and investigations. We analyzed ChatGPT’s communication quality through 14 qualitative metrics by computing patient data at four different levels to act as prompts. These included patient history, drug history, examination findings, and clinical investigations.

**Results:** Our results showed that at the initial prompt, ChatGPT’s responses were comprehensive for most (8 out of 14) variables and correct but inadequate for some (3 out of 14) variables in the majority (>50.0%) of uveitis cases. *Ethical considerations* was the only variable in terms of which responses consistently showed mixed accuracy and outdated data across all prompts in most (95.8%) uveitis cases. Also, none of the ChatGPT responses were completely inaccurate in terms of any variable at any prompt for any uveitis case.

**Conclusion:** The results reveal ChatGPT’s strengths and limitations in answering queries for patients with uveitis or its differential diagnosis while emphasizing the indispensable role of physicians in ethical decision-making.

## Introduction

The ever-expanding sphere of artificial intelligence (AI) has transformed the digital life around us. The development of large language models (LLMs) has been particularly notable, from OpenAI’s Generative Pretrained Transformer (GPT) to competing models like Google’s Gemini, Anthropic’s Claude, and Meta’s Llama(1–3). These LLMs can understand the context of each question and generate human-like responses across various domains. AI has also seamlessly integrated itself into the field of healthcare and introduced new dimensions to patient guidance(4) and physician insight(5). Although GPT-3 was not originally developed for use in medicine, its capabilities as an LLM have shown significant promise(6). In fact, ChatGPT, an LLM prototype of GPT-3, has demonstrated over 80% accuracy in suggesting differentials for common clinical diagnoses(7). Current research suggests that that ophthalmic patients are now increasingly likely to utilize AI algorithms, often chatbots, to discuss their immediate symptoms and signs prior to a clinical appointment, allowing for potential triage to optimize healthcare system flow(8,9).

While there have been positive outcomes about its incorporation into clinical practice, the completeness of the conclusions drawn by ChatGPT while answering questions is still unexplored. This especially holds true for more intricate medical conditions, including ocular conditions like uveitis. As one of the leading causes of blindness affecting all age groups globally, uveitis is a challenging diagnosis and often has a complex presentation(10,11). Additionally, patients often incur delayed management and suboptimal outcomes owing to the dearth of evidence-based guidelines(12) and scarcity of uveitis specialists worldwide(13). Hence, attempts to fill the gaps with technological advancements are now being made as patients often resort to online resources as the first point of consult(14). Since AI chatbots are now leading the game of answering patient queries, recent studies have explored ChatGPT’s capabilities in uveitis diagnosis, demonstrating AI accuracy rates comparable to that of uveitis specialists(15,16).

With the initial benchmarks for AI performance in uveitis diagnosis set forth, our proof-of-concept research aims to gather unique insights by examining the completeness and quality of AI-generated communication on real-world uveitis presentations. We hypothesize that ChatGPT, among other evolving AI models, provides reasonable explanations to uveitis patients and guides them in the right direction of ophthalmic care, in addition to assisting ophthalmologists, especially those without formal uveitis training or those in remote areas, to effectively manage complex cases. This is to ensure timely diagnoses and treatment to reduce uveitis-associated blindness rates globally. Our results would also be crucial for understanding AI’s potential in supporting ophthalmic physicians and patients to help mitigate the increasing burden of vision-threatening morbidities on healthcare resources.

## Methods

### Data Collection and Processing

We developed a robust methodology, starting with the collection of a diverse set of real-world acute uveitis cases from medical records of patients seen at a tertiary care hospital with a confirmed diagnosis. Owing to the rarity of the condition and pilot nature of our study, only 24 cases representing various uveitic entities and complexities were selected by an expert from the electronic medical records (EMR). Each case was chosen based on its completeness of documentation and final diagnosis confirmation. All 24 cases of acute uveitis had thus been diagnosed, investigated, and treated by uveitis specialists. After noting the chief complaints, past medical, surgical, ocular, family, and social history, baseline slit lamp examination, clinical investigations (hematological, radiological and ocular where relevant), and treatment advised by the specialist, the case charts for each patient were prepared based on graded details for AI prompting. After extracting patient presentations, the data was reviewed and processed to remove any unique identifiers or protected patient health information (PHI) to maintain patient anonymity. The attending’s assessment and plan were also collected and recorded as the gold standard or benchmark for the AI to be compared against. ***Figure 1*** illustrates the flowchart of data collection and processing for unique uveitis case creation.

**Figure 1:**
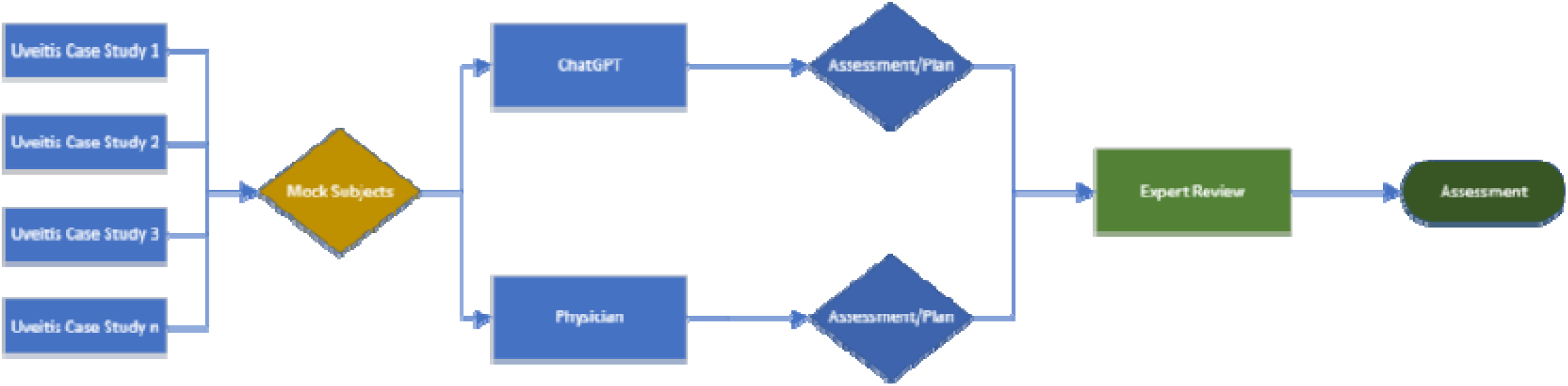
Flowchart of data collection and process for uveitis assessment.

### Experimental Design

We tested the GPT-3.5 model, (OpenAI, August 2023 release, California, USA) in this study. We used the same model and version for each of the uveitis cases to ensure uniformity and standardization in the study. Each presentation, as documented in EMR, was provided separately to the program after being de-identified to protect patient data. Our experimental design used a standardized prompt in addition to patient case findings to guide the AI’s output. The standardized prompt skeleton (SPS) read as follows: ‘In the context of the patient’s medical history, medication and examination [above/below], please formulate an assessment to explore potential causes. Additionally, outline a comprehensive plan for diagnostic investigations and initial management strategies.’ To evaluate the model’s performance with incomplete data, we tested four different styles of case presentation for each patient, with each subsequent prompt adding more information to the vignette provided previously as illustrated in ***Figure 2***. These prompts were as follows:

**Figure 2:**
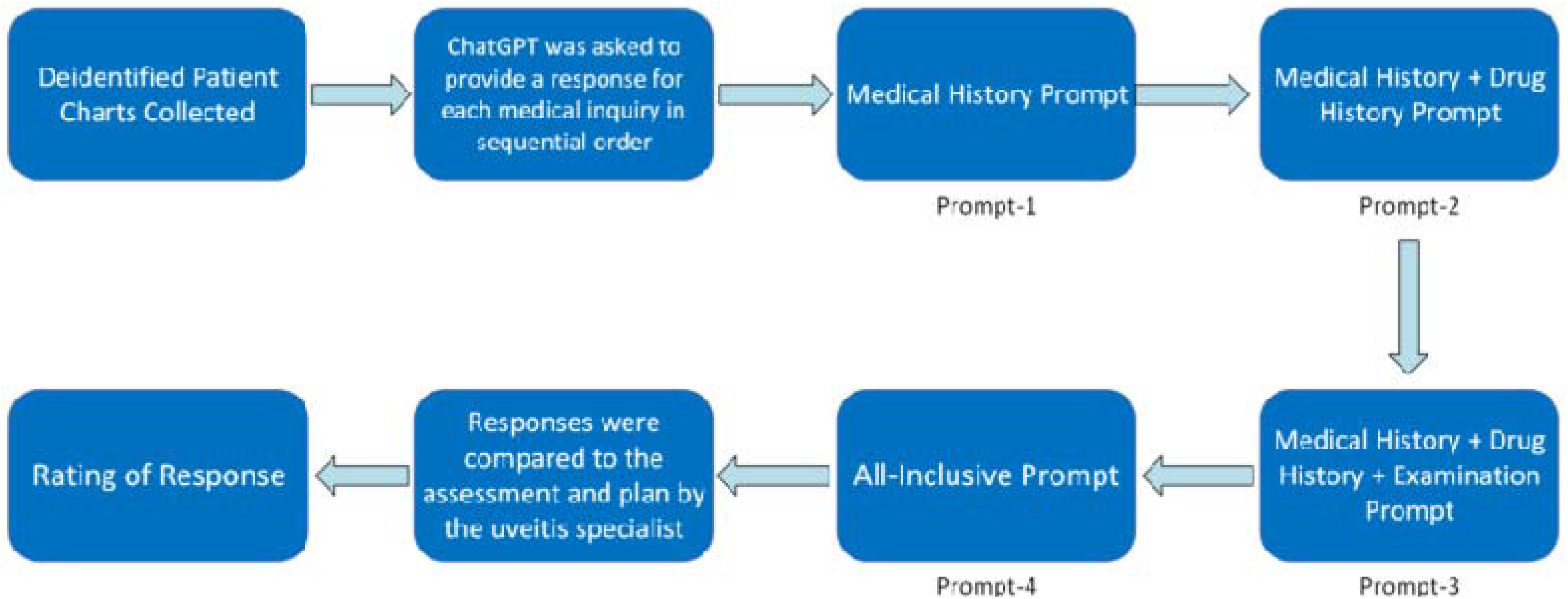
Flowchart depicting four levels of prompts.

- P1: Patient history
- P2: Patient history + Drug history
- P3: Patient history + Drug history + Physical examination findings
- P4: Patient history + Drug history + Physical examination findings + Clinical investigations

We randomized the order of case input using a computer program to mitigate order bias. Each response by ChatGPT was collected twice and copied onto an online spreadsheet for the rest of the study process. The details for the experimental design are provided in ***Figure 3***.

**Figure 3:**
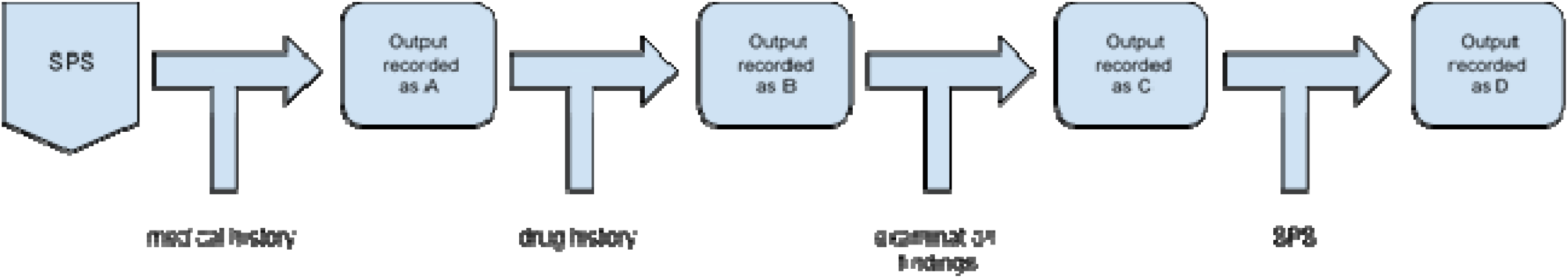
Flowchart of experimental design for uveitis assessment.

### Data Analysis

We assessed ChatGPT (GPT-3.5) output on qualitative metrics as determined by the study’s principal investigator:

- *Clinical judgment:* Evaluation of the clinical judgment demonstrated in responses. This metric considered how well both physicians and ChatGPT recognized and interpreted clinical information to provide appropriate recommendations or plans.
- *Contextual understanding:* The ability of responses to understand the context of the patient’s medical history and present concerns. The responses were assessed to evaluate how well ChatGPT tailored the answers to specific patient scenarios.
- *Nuanced responses:* The nuances and subtleties that are often present in medical cases. The depth of understanding demonstrated in identifying rare conditions or complex diagnostic considerations was assessed.
- *Patient-centric approach:* Elements such as empathy, bedside manner, and the ability to address patient concerns beyond clinical facts were assessed for this metric.
- *Cohesiveness and organization:* The structure and organization of responses were evaluated. Factors such as whether the information was well-presented, logically structured, and easy for a layperson to follow were assessed.
- *Explanation of medical concepts:* The quality of explanation of medical concepts was assessed. Whether ChatGPT provided clear explanations that were understandable to a patient, or a non-medical audience was considered.
- *Risk-benefit discussions:* Whether the responses adequately discussed the risks and benefits of proposed interventions was evaluated. This metric reflects the thoroughness of the information provided.
- *Ethical considerations:* The inclusion of ethical considerations in the responses, such as patient autonomy, informed consent, and potential conflicts of interest was assessed.
- *Tailored recommendations:* How well ChatGPT provided recommendations that were tailored to the patient’s specific medical history, preferences, and needs was evaluated.
- *Handling of uncertainty:* How the response handled uncertainty was considered. ChatGPT’s ability to acknowledge insufficient information and provide appropriate next steps was assessed.
- *Use of evidence:* Whether responses referenced evidence-based guidelines, research, or clinical studies was evaluated. This metric indicated the level of reliance on established medical knowledge.
- *Communication style:* The communication style of responses, clarity, conciseness, and the use of language appropriate for the audience was assessed.
- *Awareness of limitations:* Whether ChatGPT was aware of its own limitations was assessed. This included ChatGPT’s acknowledgment that it is an AI language model and that the cases might require specialist input.
- *Inclusion of red flags:* Whether the responses included warning signs or red flags that would necessitate urgent medical attention was evaluated.

Each of the above metrics was scored using a four-point scale as below:

- Comprehensive (Score: 4/4)
- Correct but inadequate (Score: 3/4)
- Mixed with correct/incorrect and outdated data (Score: 2/4)
- Completely incorrect (Score: 1/4)

All the qualitative metrics were scored using whole integers with no partial points allowed. Descriptive statistics were used to analyze ChatGPT’s performance, calculating the frequency and percentage of responses for each score category across the 14 qualitative metrics. This analysis was applied to evaluate performance across different levels of information provided in the prompts. Response on each prompt was collected twice to ensure the credibility of ChatGPT 3.5.

### Ethical Considerations

The study was conducted in accordance with the Declaration of Helsinki. Approval of retrospective studies was obtained from the IRB committee of Stanford University (IRB-68008). All data was anonymized and maintained with confidentiality. Stringent measures were implemented to protect patient privacy. All potential ethical concerns related to AI involvement in healthcare were considered.

## Results

Our results, as summarized in ***Figure 4*** and ***Table 1***, showed that ChatGPT’s responses were comprehensive for *communication style* (100.0%), *explanation of medical concepts* (95.8%), *cohesiveness and organization* (95.8%), *patient-centric approach* (87.5%), *contextual understanding* (79.2%), *nuanced responses* (66.7%), *clinical judgement* (62.5%), and *tailored recommendation* (58.3%) in most uveitis cases at the initial prompt. The response accuracy also generally improved for these variables when additional patient information was prompted. However, ChatGPT’s response accuracy was subpar in terms of its *use of evidence* (25.0%), *handling uncertainty* (16.7%), *risk-benefit discussions* (4.2%), *ethical considerations* (4.2%), *inclusion of red flags* (4.2%), and *awareness of limitations* (0.0%) for uveitis diagnosis at the initial prompt. Amongst these variables, response completeness only improved for *use of evidence* (25.0% at P1 to 37.5% at P4) while other variables showed a worsening or stagnant performance with additional prompts. Notably, ChatGPT’s responses in terms of *ethical considerations* were consistently mixed with correct/incorrect and outdated data in 95.8% of uveitis cases regardless of prompts. However, none of the responses were completely inaccurate in terms of any variable for any uveitis case at any prompt.

**Table 1:** Tabulated results showing diagnostic accuracy of ChatGPT for variables at each prompt.

| VARIABLES | Comprehensive (%) |  |  |  | Correct But Inadequate (%) |  |  |  |
| --- | --- | --- | --- | --- | --- | --- | --- | --- |
|  | P1 | P2 | P3 | P4 | P1 | P2 | P3 | P4 |
| Clinical Judgment | 62.5 | 50.0 | 54.2 | 62.5 | 37.5 | 50.0 | 45.8 | 37.5 |
| Cohesiveness and Organization | 95.8 | 100.0 | 100.0 | 100.0 | 4.2 | 0.0 | 0.0 | 0.0 |
| Contextual Understanding | 79.2 | 66.7 | 70.8 | 79.2 | 20.8 | 29.2 | 29.2 | 20.8 |
| Nuanced Responses | 66.7 | 62.5 | 66.7 | 75.0 | 33.3 | 37.5 | 33.3 | 25.0 |
| Patient-Centric Approach | 87.5 | 100.0 | 100.0 | 100.0 | 12.5 | 0.0 | 0.0 | 0.0 |
| Use of Evidence | 25.0 | 29.2 | 29.2 | 37.5 | 70.8 | 66.7 | 70.8 | 62.5 |
| Explanation of medical concepts | 95.8 | 100.0 | 95.8 | 100.0 | 4.2 | 0.0 | 4.2 | 0.0 |
| Risk-Benefit Discussions | 4.2 | 0.0 | 0.0 | 0.0 | 91.7 | 95.8 | 100.0 | 100.0 |
| Ethical Considerations | 4.2 | 0.0 | 0.0 | 0.0 | 0.0 | 4.2 | 4.2 | 4.2 |
| Tailored Recommendation | 58.3 | 50.0 | 58.3 | 66.7 | 41.7 | 50.0 | 41.7 | 33.3 |
| Handling Uncertainty | 16.7 | 16.7 | 12.5 | 16.7 | 83.3 | 83.3 | 87.5 | 83.3 |
| Awareness of Limitations | 0.0 | 4.2 | 4.2 | 0.0 | 100.0 | 95.8 | 95.8 | 100.0 |
| Communication Style | 100.0 | 100.0 | 100.0 | 100.0 | 0.0 | 0.0 | 0.0 | 0.0 |
| Inclusion of Red Flags | 4.2 | 8.3 | 8.3 | 4.2 | 95.8 | 87.5 | 91.7 | 95.8 |
| VARIABLES | Mixed with correct/incorrect and outdated data (%) |  |  |  | Completely Inaccurate (%) |  |  |  |
|  | P1 | P2 | P3 | P4 | P1 | P2 | P3 | P4 |
| Clinical Judgment | 0.0 | 0.0 | 0.0 | 0.0 | 0.0 | 0.0 | 0.0 | 0.0 |
| Cohesiveness and Organization | 0.0 | 0.0 | 0.0 | 0.0 | 0.0 | 0.0 | 0.0 | 0.0 |
| Contextual Understanding | 0.0 | 4.2 | 0.0 | 0.0 | 0.0 | 0.0 | 0.0 | 0.0 |
| Nuanced Responses | 0.0 | 0.0 | 0.0 | 0.0 | 0.0 | 0.0 | 0.0 | 0.0 |
| Patient-Centric Approach | 0.0 | 0.0 | 0.0 | 0.0 | 0.0 | 0.0 | 0.0 | 0.0 |
| Use of Evidence | 4.2 | 4.2 | 0.0 | 0.0 | 0.0 | 0.0 | 0.0 | 0.0 |
| Explanation of medical concepts | 0.0 | 0.0 | 0.0 | 0.0 | 0.0 | 0.0 | 0.0 | 0.0 |
| Risk-Benefit Discussions | 4.2 | 4.2 | 0.0 | 0.0 | 0.0 | 0.0 | 0.0 | 0.0 |
| Ethical Considerations | 95.8 | 95.8 | 95.8 | 95.8 | 0.0 | 0.0 | 0.0 | 0.0 |
| Tailored Recommendation | 0.0 | 0.0 | 0.0 | 0.0 | 0.0 | 0.0 | 0.0 | 0.0 |
| Handling Uncertainty | 0.0 | 0.0 | 0.0 | 0.0 | 0.0 | 0.0 | 0.0 | 0.0 |
| Awareness of Limitations | 0.0 | 0.0 | 0.0 | 0.0 | 0.0 | 0.0 | 0.0 | 0.0 |
| Communication Style | 0.0 | 0.0 | 0.0 | 0.0 | 0.0 | 0.0 | 0.0 | 0.0 |
| Inclusion of Red Flags | 0.0 | 4.2 | 0.0 | 0.0 | 0.0 | 0.0 | 0.0 | 0.0 |

**Figure 4:**
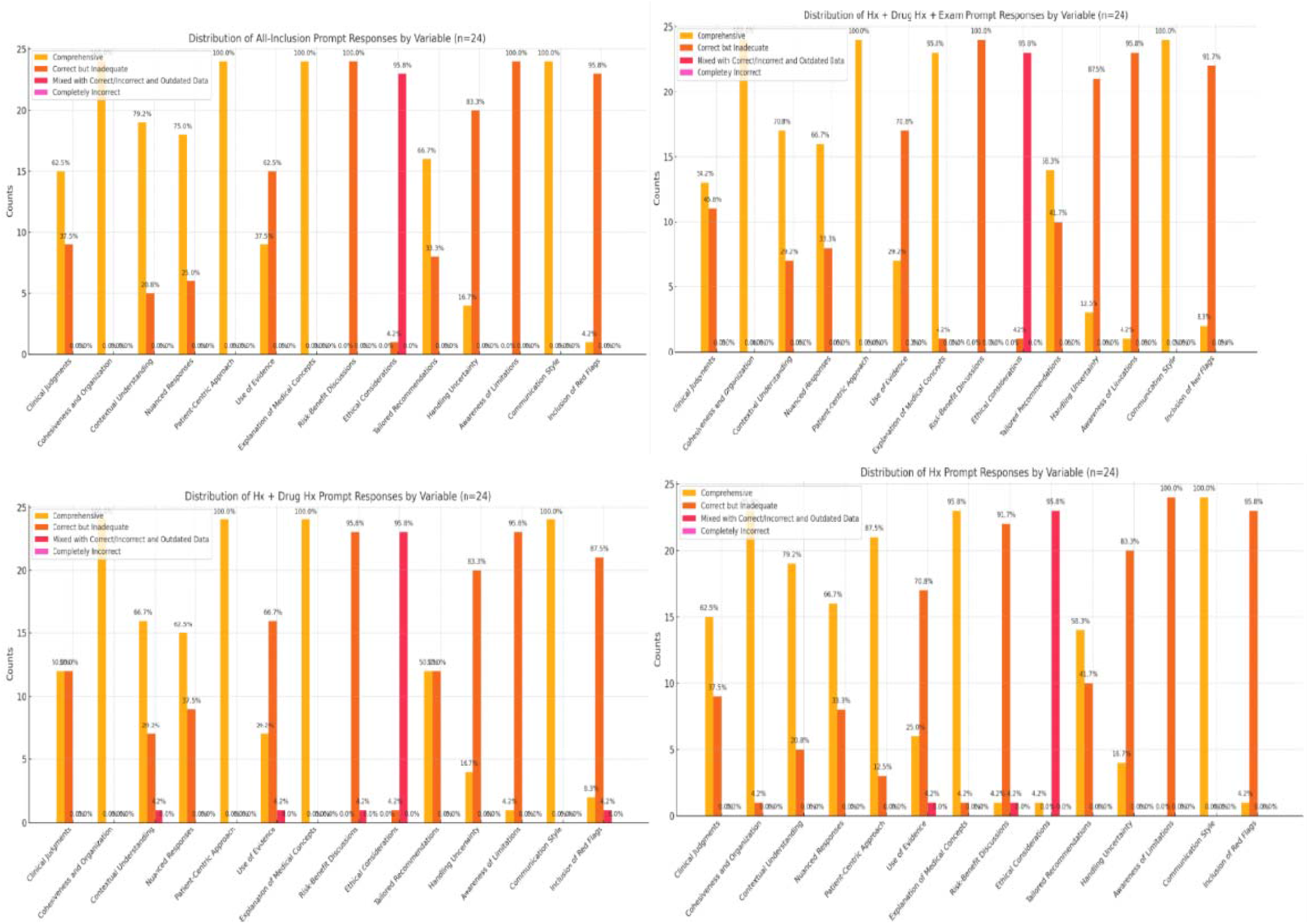
Graphs showing the diagnostic accuracy of ChatGPT responses at each level of prompt.

ChatGPT responses showed incredible performance in terms of *patient-centric approach*, with 87.5% of cases showing accuracy at the initial prompt which scaled up to 100% with the subsequent prompts. Similarly, in terms of *cohesiveness and organization*, ChatGPT responses were complete for 95.8% of cases at the initial prompt which increased and stabilized to 100% accuracy with additional prompts. *Communication style* was also a strongpoint of ChatGPT responses with 100% comprehensiveness for all uveitis cases across all prompts. Lastly, the *explanation of medical concepts* was also well done by ChatGPT for all uveitis case presentations, albeit with a few (4.2%) responses showing correct but inadequate information at P2 and P4.

With respect to *clinical judgment*, 62.5% of the responses generated were completely accurate while the rest 37.5% were correct but lacked information at the initial prompt. With additional information provided, comprehensiveness increased from 50.0% at P2 to 54.2% at P3 but stabilized to 62.5% at the final prompt. While the information shared by ChatGPT may be inadequate for this variable, no uveitis case prompted responses with mixed accuracy or complete inaccuracy. Regarding *contextual understanding*, the comprehensiveness of ChatGPT responses scaled down from 79.2% at P1 to 66.7% at P2 but increased to 70.8% at P3 and readjusted to 79.2% at P4. The remaining responses were correct but inadequate at each prompt, albeit 4.2% cases showing mixed accuracy at P2. Similarly, *nuanced responses* were initially comprehensive for 66.7% of uveitis cases but fluctuated, i.e. down to 62.5% at P2, back to 66.7% at P3, and up to 75.0% at P4, with the remaining responses being correct but inadequate at each prompt.

The *use of evidence* was suboptimal in most ChatGPT responses with 70.8% of uveitis cases having correct but inadequate information while 4.2% responses had mixed accuracy at the initial prompt. However, the comprehensiveness increased incrementally from 25.0% at P1 to 37.5% at P4. *Risk-benefit discussions* consistently showed high inadequacy in the data which increased with each prompt, i.e. 91.7% cases showing correct but incomplete information at P1 scaling up to`100.0% at P4. ChatGPT responses to uveitis management also performed poorly in term of *ethical considerations*, with 95.8% of responses showing mixed accuracy and outdated data across all prompts, and the comprehensiveness reduced from 4.2% to 0.0% with the addition of drug history at P2. ChatGPT was also ill-equipped in terms of generating responses *handling uncertainty* for uveitis cases, with comprehensiveness equaling 16.7% at all prompts, albeit at P3 when it reduced further to 12.5%, with the remaining responses being correct but incomplete. The *awareness of limitations* was also one of the shortcomings of ChatGPT toward uveitis management as the responses demonstrated high inadequacy in the information presented, and comprehensiveness only mildly increased from 0.0% to 4.2% with the addition of drug history (P2) and exam findings (P3) but reduced back to 0.0% at P4. Lastly, ChatGPT was also limited in its response comprehensiveness by the *inclusion of red flags* since most cases generated accurate but incomplete information across all prompts, with some content (4.2%) showing mixed accuracy and outdated data with drug history addition at P2.

## Discussion

Our findings demonstrate the promising potential of ChatGPT in summarizing case details, generating initial differentials, and providing patient-centric explanations for most uveitis presentations, albeit with some inaccuracies when considering ethicalities, uncertainties, and emergencies. These results are in congruence with previous findings where ChatGPT provided understandable and relevant suggestions to improve clinical decision-making for healthcare workers(17). Since ChatGPT also explained medical concepts in a well-organized and cohesive manner, it can potentially serve as a valuable tool to answer patients’ queries and accurately guide them towards visiting a uveitis specialist when warranted by their symptoms as concurred by a recent similar study(18). ChatGPT also showed impressive accuracy and consistency in clinical judgement of uveitis cases, further confirming their utility to assist physicians, particularly ophthalmologists, in devising evidence- and case-based care as also proven by Chen et. al.(19).

However, responses by ChatGPT served low adequacy in terms of discussing risk-benefits, red flags, and limitations for uveitis cases despite prompting additional patient information in our study. This indicates that while ChatGPT may be excellent in giving clinically comprehensive responses, it lacks in suggesting improved outcomes in medicolegal and ethical aspects, which sustain as areas better handled by physicians. While AI chatbots could be potentially useful tools to assist clinicians in their decision-making processes, their performance may not yet be at the same level as experienced clinicians for deployment. Through comprehensive ethical guidelines, ophthalmologists can thus ensure the responsible use of ChatGPT by promoting reliable information exchange, protecting patient privacy, and empowering uveitis patients to make informed decisions about their health(20).

Our results also showed that response completeness fluctuated with additional prompts for some variables. Possible reasons for this variability, as cited in ophthalmic literature, include inconsistencies in training data, sensitivity to prompt information, random output generation, or limitations in maintaining context across queries(21,22). The high proportion of responses mixed with inaccurate or outdated data as found in our study has also been previously recognized as ‘AI hallucinations’ and defined as invented information by the chatbot causing a serious challenge for its use in medical practice(23). This variability is thus proof that despite its great promise and vast expanse of knowledge, AI models can only be justified as a supplement rather than a substitute to healthcare professionals(24).

This study has several limitations. Potential confounding factors include: (a) the ‘gold standard’ set forth in our study being the attending’s plan, which may differ for some aspects of real-world management across different physicians, (b) variation in how ChatGPT handles changes in prompting structure from other LLMs, including newer paid versions of ChatGPT, which were never tested in our study, (c) potential selection bias owing to the uveitis cases shortlisted by a single expert from our team which might be skewed toward certain complexities or uveitis types, (d) the inability of GPT-3.5 to incorporate external references or recent literature unless specifically quoted in the prompts, and (e) the non-replicability of the generated responses using health records if patient data differs in terms of demographic or comorbid variables.

## Conclusion

In conclusion, our proof-of-concept study provides insights into the evolving relationship between AI and ophthalmic care and the need for cautious integration and continuous improvement in specialized medical domains. AI chatbots such as ChatGPT indeed have potential in educating patients, addressing the shortage of uveitis specialists in developing countries, and serving as an aid to clinicians regarding uveitis diagnoses and treatment(25). Fixing inconsistencies within variables identified by our study may thus necessitate ongoing human oversight, increased dataset size, rigorous training, comprehensive testing, and further validation of the AI models to better complement ophthalmic care.

## Data Availability

All data produced in the present study are available upon reasonable request to the authors.

## Disclosures

### Authors’ Contributions

MSH and YJS conceptualized the study. YJS supervised the study. AHK and AK collected the data. TJ and KJ analyzed the data. MSH, AHK, ZZF and ANTT drafted the initial manuscript. AHK and ZZF contributed equally to making the final edits. All authors reviewed and approved the final draft.

### Conflict of Interest

The authors have no conflict of interests to declare.

### Funding

This project received in-kind support from the National Eye Institute (P30EY026877) as part of the Stanford Vision Research Core Award for the Byers Eye Institute at Stanford Medicine.

## Acknowledgments

None.

## Notes

### Competing Interest Statement

The authors have declared no competing interest.

### Summary of Updates

Funding statement was corrected. No major changes to the manuscript content or methods otherwise.

